# Updated Health Opportunity Cost Estimates for 92 Low- and Middle-Income Countries: Implications for Global Health Financing and Donor Allocation

**DOI:** 10.64898/2026.03.31.26349880

**Authors:** Jessica Ochalek

## Abstract

**Background:** Estimates of the marginal cost per disability-adjusted life year (DALY) averted from government health expenditure (GHE) provide an empirical basis for allocating scarce health resources to maximise population health. Existing cross-country estimates have informed priority setting in several countries and international policy discussions but are based on data that are now more than a decade old, during which patterns of health expenditure, disease burden, and global health financing have changed substantially.

**Methods:** This paper provides updated estimates of the marginal cost per DALY averted for 92 low- and middle-income countries (LMIC), applying previously estimated elasticities of the effect of GHE on health outcomes from Ochalek et al. (2018) to recent mortality, morbidity, population, and GHE data. We model two scenarios: (1) countries raising domestic health expenditure to a tiered minimum of 10% or 15% of general government expenditure, depending on current level, with Abuja Declaration-consistent results for African Union countries reported separately; and (2) reallocating development assistance for health (DAH), considering both the targeting of existing on-budget support and the potential of the full DAH envelope. We use the estimated elasticities to reflect diminishing marginal returns to health expenditure when calculating the health gains associated with additional resources.

**Results:** Updated estimates of the marginal costs per DALY averted range from approximately $54 to $18,547 across countries (average across DALY measures). In most countries (68%), estimates are higher than in the previous analysis, largely reflecting differences in GHE. Increasing domestic expenditure to the tiered minimum would avert 145 million DALYs but require $392 billion across countries. Reallocating the $11.2 billion in on-budget DAH according to country-specific opportunity costs would avert 75.3 million DALYs compared to the estimated 45.3 million averted by the current allocation of on-budget spending, though this requires redirecting funding away from many currently-funded countries. There is further potential to avert 66.1 million DALYs by optimally allocating the 25.7 billion portion of the DAH envelope that comes from off-budget, NGO and philanthropic resources, measured against a baseline in which they avert no DALYs.

**Conclusion:** Updated estimates provide an empirical basis for informing both domestic priority setting and the allocation of international health financing. Aligning donor funding with country-specific opportunity costs could substantially increase the global health gains achieved with limited resources.

## Background

All health systems face resource constraints and difficult trade-offs about what healthcare to provide with pooled government funds. Every government dollar spent on health carries an opportunity cost: financing one intervention inevitably displaces another which would also have generated health. These constraints are typically more binding, and the trade-offs more consequential, in low- and middle-income countries (LMIC), where fiscal space is limited, disease burdens are higher, and many highly cost-effective essential services remain underfunded. The situation has become more acute amid tightening fiscal conditions and declining external support. External aid for health fell 21% between 2024 and 2025 to $39.1 billion and is projected to continue to decline to $36.2 billion by 2030 (1,2). 1.38 billion people around the world are living in LMIC which are both highly exposed to US global health aid cuts and highly fiscally constrained (3).

Country-specific estimates of the marginal cost per DALY averted from government health expenditure (GHE) are essential for informing priority setting within health systems. These estimates reflect the health that a marginal increase in expenditure would generate (or equivalently, the health that would be forgone from a marginal reduction in expenditure). These estimates can be used to provide a benchmark (e.g., a cost-effectiveness threshold) for assessing whether new interventions or programmes are likely to increase or reduce population health (4). They can also be used to quantify the extent to which interventions or programmes will increase or reduce population health (i.e., net health effect) (5,6). This type of analysis has been used to inform health benefits package design in Malawi and Uganda (7,8) and decisions around the expansion or contraction of bundles of services in Ghana (9).

It can also be used to inform the health consequences of broader changes in GHE (10). One potential response to cuts in donor funding would be for countries to increase domestic funding for healthcare. Such increases were already a policy objective for countries under the Abuja Declaration in 2001, where they pledged to allocate at least 15% of their annual budget to GHE (11). As of writing, however, well over half of LMIC have yet to meet this target. Estimates of the marginal cost per DALY averted can inform the health effects of expanding GHE.

Such estimates are also important for informing donor resource allocation decisions around what programmes to invest in and in which healthcare systems. Amid the backdrop of cuts to health aid, there is renewed urgency for methods that can guide more effective donor engagement and align external financing with national priorities. One prominent reform proposal is the “New Compact” for global health financing, which seeks to rebalance responsibilities between governments and donors. The New Compact is based on three pillars that reflect a shared responsibility between a country and its donors: (1) evidence-informed, locally led prioritization; (2) domestic-first resource allocation, with donor support for interventions that are only marginally cost-effective at the country level; and (3) consolidated supplementary aid (12–14). By shifting donors away from earmarked disease-specific funding toward supporting marginal expansions of nationally determined service packages, the New Compact implies a need to consider opportunity costs and the marginal productivity of health expenditure when making financing decisions. Operationalising such an approach requires estimates of the marginal productivity of health expenditure across countries reflecting health opportunity costs in each country. Without this empirical foundation, calls for domestic-first financing and consolidated supplementary aid remain conceptually appealing but difficult to implement in practice.

The opportunity cost of expenditure on a given intervention for a specific disease or population falls across the population that benefits from healthcare expenditure. Therefore, consideration of opportunity cost requires a common measure of health that enables comparison across the range of services and interventions that may be provided by the healthcare system. The disability-adjusted life year (DALY) is an example of such a measure by capturing both premature mortality and time lived with disability. Expressed as DALYs averted, health gains from interventions that extend life, improve quality of life, or both can be compared on a common scale, enabling prioritisation across the full range of health services within a national package. Another measure that has been used is quality-adjusted life years (QALYs), which capture changes in both survival and health-related quality of life and are widely used in health technology assessment in high-income countries. Estimating QALYs requires preference-based health state value sets (e.g., EQ-5D tariffs), which have historically been unavailable in many LMIC (15). Estimates of the marginal cost per DALY averted or QALY gained from GHE are currently available for only a small number of countries (13 at the time of writing) (16). Estimation is data intensive and can take years (17). A 2018 study by Ochalek et al. provided cross-country estimates of the marginal productivity of health expenditure and corresponding estimates of the marginal cost per DALY averted (18). However, the global health financing landscape has changed over the last decade. Patterns of health expenditure have shifted, disease burdens have evolved, demographic structures have aged in many settings, and donor funding levels have become more volatile. Up to date estimates are required to better inform domestic and international decision-making around allocating funding for healthcare.

This paper provides updated estimates of the marginal cost per DALY averted and uses these to model the potential responses to changes in the donor funding landscape. We first examine the implications of governments increasing domestic health expenditure to a tiered minimum of 10% or 15% of general government expenditure, depending on their current level. We then assess the health effects of allocating DAH according to New Compact principles, considering both the reallocation of existing on-budget support and the potential of the full DAH envelope if optimally targeted.

## Methods

We update country-specific estimates of the marginal cost per DALY averted using the health opportunity cost framework developed by Ochalek et al. (2018) (19). Rather than re-estimating the elasticity of health outcomes with respect to GHE, we retain the previously estimated central elasticity parameter and update the underlying country-level inputs. This approach assumes that the proportional responsiveness of health outcomes to expenditure remains stable over time, while allowing baseline levels of expenditure, disease burden, and population to evolve.

Following Ochalek et al. (2018), we construct four alternative DALY-based measures (DALYs 1–4) to ensure consistency with the underlying elasticity estimates. The first measure is grounded in mortality data and uses the estimated elasticity of mortality with respect to expenditure to calculate DALYs averted. The second measure relies on the elasticity of survival (years of life lost, YLLs) with respect to expenditure. The third measure combines the elasticities of survival and morbidity (years lived with disability, YLDs), applying each to the corresponding burden component. The fourth measure applies the elasticity estimated directly for DALYs. For each measure, the relevant elasticity is applied to the updated country-level outcome data to estimate DALYs averted. This results in four estimates of DALYs averted and we also calculate the average DALYs averted across the four estimates for presentational purposes.

Cost per DALY averted is calculated from a 1% proportional increase in GHE. GHE is defined as spending through government schemes, including tax-based financing, compulsory contributory health insurance schemes and on budget donor support. For each country, the estimated percentage change in the relevant health outcome associated with a 1% expenditure increase is converted into absolute DALYs averted using updated healthcare expenditure, demographic, mortality and morbidity data, and the marginal cost per DALY averted is calculated.

We present results for each country both as an absolute cost per DALY averted and as a proportion of 1x and 0.5x GDP per capita. Expressing estimates relative to GDP per capita facilitates comparison with historically used cost-effectiveness benchmarks, which, despite well-documented limitations, remain influential in policy discussions and global health decision-making (e.g., influencing cost-effectiveness analysis in Disease Control Priorities, Third Edition) (4,20–22).

Such estimates are appropriate for calculating health effects of marginal changes in expenditure, but they do not reveal the relationship between health and expenditure for more substantial quantities. Consistent with the underlying econometrics, which estimates the effect as an elasticity, there are likely diminishing marginal returns in this relationship. This means that the absolute cost per DALY at the margin is increasing with expenditure (10).

We adopt the approach outlined in Lomas et al. (2025), which enables us to evaluate two separate illustrative applications: increases in domestic health expenditure and reallocation of external health financing across countries. These scenarios answer different policy questions and are not intended to represent a fully specified implementation of the New Compact, but to illustrate the implications of two elements central to it: increased domestic financing and supplementary donor allocation informed by country-specific opportunity costs. For ease of exposition, we base our analysis on DALY 4 since this is based on one estimated elasticity. Since DALY 4 is at the upper or lower end of the range of estimates from DALYs 1-4 in only 25 of 92 countries, results based on this should be regarded as broadly representative of our results more generally.

To illustrate the potential health effects of increased domestic financing, we modelled a stepped increase in the share of general government expenditure (GGE) allocated to health. Countries allocating less than 10% of GGE to health were assumed to increase this share to 10%; countries allocating between 10% and less than 15% were assumed to increase it to 15%; and no increase was assumed for countries already allocating at least 15%. 15% has been set as a target for African Union member state governments in the Abuja Declaration (11), but for many countries it represents a large and potentially unrealistic increase from current spending levels. As such, we apply lower thresholds to illustrate more incremental increases in domestic financing. However, these thresholds are intended as illustrative benchmarks rather than normative recommendations. Given that 15% is a policy target for African Union member states, however, we also present results for African Union member state governments allocating 15% of national budgets to health as per the Abuja Declaration in the appendix. Ukraine is excluded from the scenario analyses, because data on general government expenditure and on the split between on- and off-budget support are not available for recent years.

Domestic GGE per capita (current US$) was calculated using data on domestic general GHE per capita and domestic general GHE as a percentage of GGE. The latter was converted to a proportion prior to calculation (see Data). For each country, the target level of health expenditure per capita consistent with the relevant 10, 15 or current % budget allocation was calculated as:

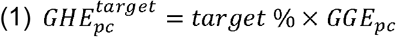

where *GGE_pc_* denotes domestic GGE per capita.

The additional expenditure required per capita was calculated as:

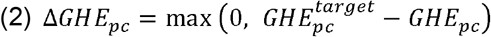

Where *GGE_pc_* denotes observed GHE per capita (as defined in the Data section). Countries already allocating at least 15% of GGE to health were assigned a value of zero additional requirement. Total additional expenditure requirements were calculated by multiplying Δ *GGE_pc_* by country population in 2023 corresponding to the year for which the most recent GHE and GGE data are available.

The number of DALYs per capita for a country can be expressed as a power function:

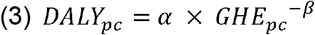

where *β* denotes the magnitude of the estimated elasticity on DALYs, *DALY_pc_* represents the DALYs per capita and *GGE_pc_* is the GHE per capita. is unknown but is solved for each country given the observed and estimated values for other parameters. We calculate the health gains of all countries achieving the objective of spending 10, 15 or current % of GGE on GHE as the difference in health gains between the original expenditure level and the objective level for countries that were spending less than that as a percent of GGE:

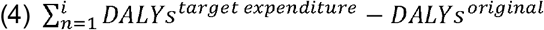

To examine the potential health gains from reallocating external health financing, we consider two distinct reallocation exercises. First, we examine whether the $11.2 billion in on-budget support currently received by countries (per WHO Global Health Expenditure Database, GHED, data) is well targeted, by comparing the DALYs averted under the current country-level allocation of this on-budget pot against a health maximising reallocation of the same $11.2 billion informed by country-specific opportunity costs. This holds the total resource envelope fixed and isolates the effect of targeting alone. Second, we examine the potential of the full DAH envelope: we remove the on-budget support component from GHE for each country (using the WHO GHED estimate), re-calculating DALYs using Equation 3, and from this baseline consider a health maximising allocation of the full IHME DAH estimate, which includes off-budget, NGO, and philanthropic channels not captured in on-budget data, across countries. Because current country-level allocation of off-budget and philanthropic funding is not comprehensively tracked, this second exercise is evaluated against a baseline of no on-budget support rather than against a defined current allocation, and should be interpreted as indicating the scale of potential impact of the total DAH envelope rather than a reallocation-efficiency result.

Total DAH (the sum of on- and off-budget support) measured via WHO GHED on-budget and off-budget data is lower than that measured by IHME. On-budget support makes up nearly half of donor funding for countries in our sample using this data. This likely reflects that funding captured in national accounts under-captures NGO-delivered programmes and vertical programmes run outside government systems. In the IHME data, approximately $10 billion of 2023 DAH flows through NGO, international NGO, and foundation channels that do not pass through recipient government budgets. For the full-DAH exercise, we apply the IHME Development Assistance for Health database estimate for 2023, which, after correcting for double-counted transfers per the IHME User Guide (23), totals $36.9 billion.^1^ This total includes an estimated $9.6 billion in private philanthropic funding and $3.9 billion in World Bank debt-repayment flows, which fall outside the scope of official development assistance; a corresponding ODA-eligible government subtotal is $23.5 billion. We use the full $36.9 billion figure for this exercise, as IHME’s broader scope, including philanthropic and NGO-delivered assistance, better reflects the full set of resources potentially available for reallocation. This is inclusive of both on and off-budget support. Obtaining the allocation of DAH that maximises health globally is achieved by iteratively re-allocating each additional dollar to the country with the lowest cost per DALY averted given the new global allocation.

## Data

Updating the estimated marginal cost per DALY averted requires country-level data on mortality and life expectancy (conditional on age at death), disaggregated by age and sex, as well as years lived with disability (YLDs). These are obtained from the most recent Global Burden of Disease (GBD) release via the Global Health Data Exchange (GHDx) (24). Consistent with the methodology described in Ochalek et al. (2018), these inputs are used to construct the four DALY measures described above.

GHE includes spending through government schemes and compulsory contributory health care financing schemes, irrespective of the original revenue source, including external funds channelled through government budgets (25). Data for this variable are drawn from the WHO GHED. WHO GHED reports on-budget external funds (transfers distributed by government from foreign origin) and off-budget direct foreign transfers. Where countries do not report this split, WHO estimates it from OECD DAC Creditor Reporting System data, treating aid channelled through the public sector as on-budget and the remainder as off-budget (26).^2^ Transfers distributed by government from foreign origin (FS.2) are reported separately in GHED and are used in this analysis to identify on-budget external support. Consistent with previous analysis, our expenditure measure excludes voluntary health care payment schemes and household out-of-pocket payments (OOPs) and off-budget donor funding (i.e., direct foreign transfers). For further discussion of the role of OOPs in health opportunity cost estimation, see Ochalek et al. (forthcoming). WHO provide additional information on per capita on- and off-budget support received by countries drawn from national health accounts in the first instance, and where this information is incomplete, surveys and reports from donors, governments, non-profit institutions may also be used (27). For Ukraine, GHED data are unavailable for 2022 and 2023, so the most recent available year (2021) is used. The Institute for Health Metrics and Evaluation (IHME) publish data on donor assistance for health (DAH) further disaggregated by source, channel of funding, and health focus and programme areas. It draws on donor project databases, financial statements, annual reports, IRS 990s, and correspondence with agencies (28). This is used to inform the full DAH envelope for 2023.

Countries are classified into income groups according to the World Bank country and lending groups using their 2025 classification, which is based on GNI per capita in 2023 (29,30).^3^

Domestic GGE per capita was not directly available for all countries and years. We therefore calculated it using reported domestic general GHE per capita and domestic general GHE as a share of total general government expenditure from WHO GHED (26). Since:

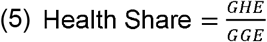

total domestic GGE per capita was calculated as:

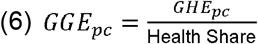

where health share is expressed as a decimal. This approach preserves consistency with national health accounts definitions and avoids mixing expenditure concepts (e.g., government final consumption expenditure).

## Results

We estimate marginal cost per DALY averted for 92 LMIC.^4^ For most countries in the analysis (68%, 63 of 92) the estimated marginal cost per DALY averted is higher in the current analysis than the previous analysis (19). Differences between the two sets of estimates closely track differences in the GHE per capita used in each analysis. GHE per capita is higher in the current analysis in 56 of 63 countries where the estimated marginal cost per DALY averted increased, and lower in 26 of the 29 countries where it decreased. In part, this reflects revisions to the GHE data rather than changes in spending over time. Using the current data for both years, GHE per capita fell between 2014 and 2023 in 15 of the 29 countries with lower estimates. Differences in changes in YLLs and total DALYs between the two groups are present but modest compared to changes in GHE.

Figure 1 gives point estimates for the cost per DALY averted from DALY 4, as this is what used for later scenario analysis. The bars show the range of cost per DALY averted estimates across the four methods of calculating DALYs averted from the data. Consistent with 2018 analysis, most (85 out of 91) LMIC for which we are able to estimate the marginal cost per DALY averted have average estimates below 1x GDP per capita.^5^

**Figure 1.**
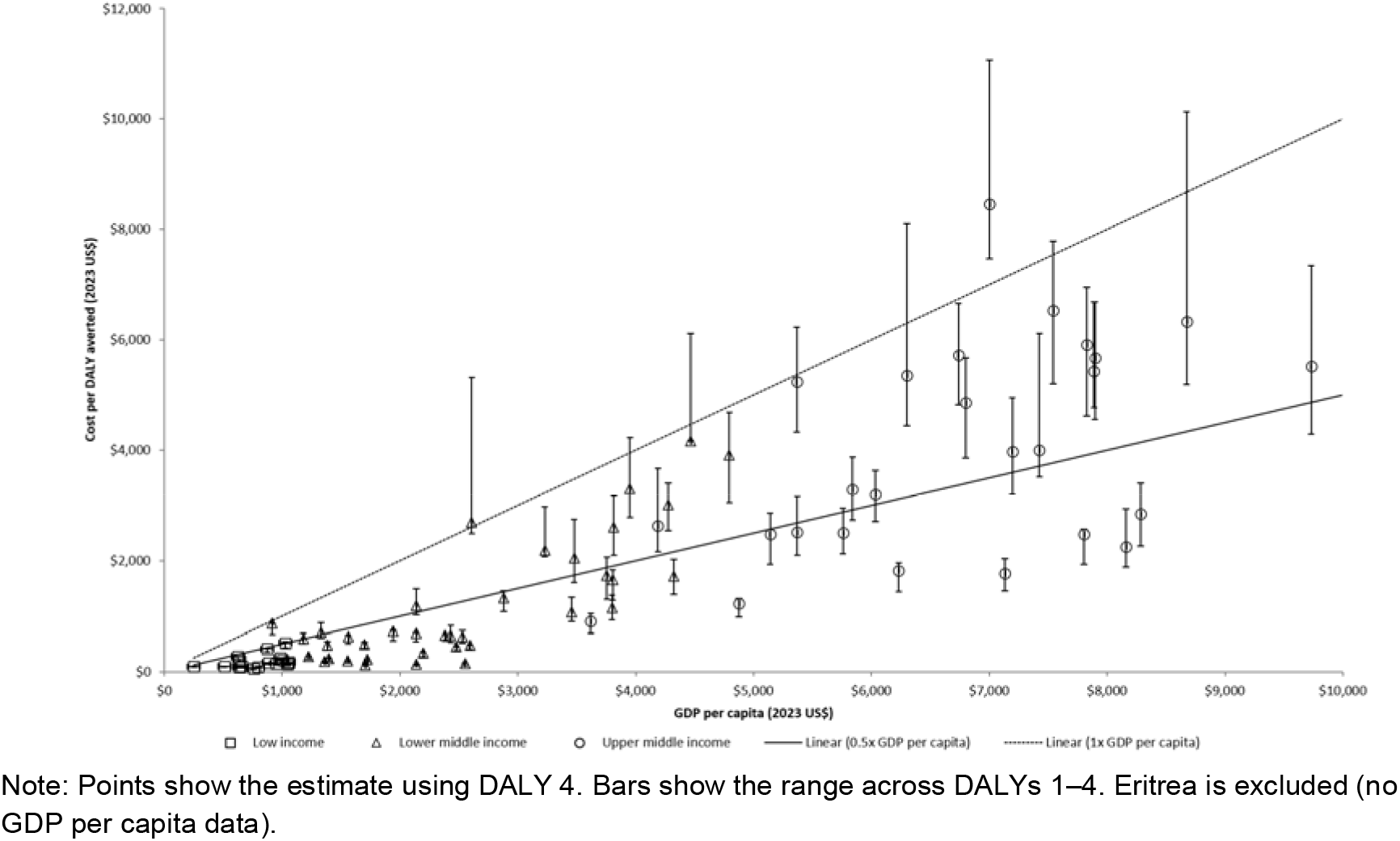
Estimated marginal cost per DALY averted from government health expenditure by country.

The estimated marginal cost per DALY averted from DALY 4 ranges from $59-520 in low-income countries, $123-4,161 in lower middle-income countries, and $903-16,852 in upper middle-income countries. Cost per DALY averted for each country from DALY 4 is reported in Table 1. (All estimates are reported in the appendix.)

**Table 1.** Estimated marginal cost per DALY averted, domestic financing requirements, and reallocation of on-budget development assistance for health, by country.

| Country | Marginal cost per DALY averted given current GHE (2023 US\$) | Increase of GHE as a % of GHE<br>If below 10% → 10%<br>10% to under 15% → 15%<br>15% or above → keeps the current % | | | | Marginal cost per DALY averted (2023 US\$) | Re-allocation of \$11.2 billion on-budget donor funding | | |
| --- | --- | --- | --- | --- | --- | --- | --- | --- | --- |
| | | Current GHE as a % of GGE | Increased GHE as a % of GGE | Additional expenditure required (millions 2023 US\$) | DALYs averted by additional expenditure (millions) | | Change in on budget funding allocated (millions 2023 US\$) | Change in DALYs averted by DAH (thousands) | Marginal cost per DALY averted (2023 US\$) |
| Albania | 5,513 | 12% | 15% | 147 | 0.02 | 6,893 | 0 | 0 | 5,513 |
| Algeria | 2,512 | 6% | 10% | 3,431 | 1.01 | 4,513 | -1 | 0 | 2,512 |
| Argentina | 16,852 | 17% | 17% | 0 | 0.00 | 16,852 | -586 | -35 | 16,620 |
| Armenia | 2,256 | 6% | 10% | 302 | 0.09 | 4,619 | -4 | -2 | 2,234 |
| Azerbaijan | 1,770 | 4% | 10% | 1,381 | 0.46 | 4,900 | -11 | -7 | 1,751 |
| Bangladesh | 143 | 2% | 10% | 4,114 | 11.40 | 779 | 753 | 3,983 | 245 |
| Belarus | 5,673 | 12% | 15% | 992 | 0.15 | 7,631 | 0 | 0 | 5,673 |
| Belize | 3,998 | 11% | 15% | 30 | 0.01 | 5,797 | -1 | 0 | 3,939 |
| Benin | 232 | 7% | 10% | 113 | 0.39 | 355 | 13 | 55 | 245 |
| Bolivia | 3,011 | 12% | 15% | 477 | 0.14 | 3,853 | -7 | -2 | 3,001 |
| Botswana | 5,914 | 15% | 15% | 0 | 0.00 | 5,914 | -35 | -6 | 5,695 |
| Brazil | 7,196 | 9% | 10% | 5,229 | 0.70 | 7,680 | -6 | -1 | 7,195 |
| Burkina Faso | 410 | 13% | 15% | 131 | 0.29 | 492 | -258 | -792 | 279 |
| Burundi | 93 | 10% | 10% | 1 | 0.01 | 94 | 152 | 980 | 245 |
| Cambodia | 651 | 8% | 10% | 198 | 0.25 | 919 | -69 | -115 | 576 |
| Cameroon | 218 | 6% | 10% | 396 | 1.33 | 399 | 62 | 268 | 245 |
| Cape Verde | 3,908 | 16% | 16% | 0 | 0.00 | 3,908 | -12 | -3 | 3,414 |
| Chad | 151 | 10% | 15% | 145 | 0.77 | 233 | 166 | 857 | 245 |
| China | 8,766 | 10% | 15% | 279,626 | 25.37 | 13,710 | -13 | -2 | 8,766 |
| Colombia | 8,457 | 18% | 18% | 0 | 0.00 | 8,457 | -4 | 0 | 8,456 |
| Comoros | 623 | 13% | 15% | 5 | 0.01 | 742 | -17 | -43 | 288 |
| Congo | 456 | 7% | 10% | 88 | 0.15 | 706 | -45 | -115 | 354 |
| Costa Rica | 16,215 | 26% | 26% | 0 | 0.00 | 16,215 | -1 | 0 | 16,212 |
| Cote d'Ivoire | 478 | 7% | 10% | 487 | 0.84 | 693 | -144 | -322 | 427 |
| Democratic<br>Republic of the<br>Congo | 93 | 7% | 10% | 262 | 2.36 | 132 | 952 | 6,151 | 245 |
| Dominican<br>Republic | 5,707 | 16% | 16% | 0 | 0.00 | 5,707 | -21 | -4 | 5,673 |
| Ecuador | 5,721 | 12% | 15% | 1,438 | 0.22 | 7,528 | -3 | -1 | 5,718 |
| Egypt | 1,078 | 8% | 10% | 1,508 | 1.21 | 1,430 | -497 | -487 | 985 |
| El Salvador | 5,240 | 19% | 19% | 0 | 0.00 | 5,240 | -4 | -1 | 5,230 |
| Eritrea | 139 | 4% | 10% | 109 | 0.46 | 379 | 51 | 274 | 245 |
| Ethiopia | 166 | 9% | 10% | 112 | 0.65 | 182 | 543 | 2,684 | 245 |
| Gabon | 2,472 | 10% | 15% | 162 | 0.05 | 3,985 | -18 | -7 | 2,343 |
| Georgia | 2,842 | 8% | 10% | 214 | 0.06 | 4,006 | -6 | -2 | 2,816 |
| Ghana | 658 | 10% | 15% | 705 | 0.84 | 1,047 | -25 | -38 | 647 |
| Guatemala | 2,501 | 18% | 18% | 0 | 0.00 | 2,501 | -107 | -44 | 2,387 |
| Guinea | 201 | 7% | 10% | 99 | 0.40 | 296 | 47 | 213 | 245 |
| Guinea-Bissau | 247 | 10% | 15% | 21 | 0.07 | 386 | 0 | -1 | 246 |
| Haiti | 123 | 9% | 10% | 11 | 0.09 | 135 | 108 | 614 | 245 |
| Honduras | 2,190 | 12% | 15% | 296 | 0.12 | 2,950 | -224 | -118 | 1,727 |
| India | 629 | 5% | 10% | 53,976 | 54.24 | 1,510 | -2,516 | -4,127 | 597 |
| Indonesia | 1,223 | 9% | 10% | 3,506 | 2.60 | 1,482 | -126 | -103 | 1,215 |
| Jamaica | 6,533 | 16% | 16% | 0 | 0.00 | 6,533 | -7 | -1 | 6,483 |
| Jordan | 4,161 | 11% | 15% | 754 | 0.15 | 5,986 | -148 | -37 | 3,874 |
| Kazakhstan | 6,339 | 11% | 15% | 2,634 | 0.34 | 9,516 | -15 | -2 | 6,324 |
| Kenya | 729 | 10% | 10% | 39 | 0.05 | 745 | -338 | -510 | 621 |
| Kyrgyzstan | 1,190 | 7% | 10% | 138 | 0.10 | 1,737 | -1 | -1 | 1,188 |
| Lebanon | 2,050 | 17% | 17% | 0 | 0.00 | 2,050 | -22 | -11 | 1,972 |
| Lesotho | 875 | 17% | 17% | 0 | 0.00 | 875 | -43 | -60 | 627 |
| Macedonia | 6,334 | 12% | 15% | 168 | 0.02 | 8,253 | 0 | 0 | 6,334 |
| Madagascar | 98 | 8% | 10% | 51 | 0.46 | 125 | 262 | 1,650 | 245 |
| Malawi | 224 | 10% | 15% | 152 | 0.54 | 347 | 27 | 115 | 245 |
| Malaysia | 5,133 | 8% | 10% | 1,979 | 0.34 | 6,762 | 0 | 0 | 5,133 |
| Mali | 137 | 6% | 10% | 197 | 1.08 | 237 | 212 | 1,146 | 245 |
| Mauritania | 698 | 8% | 10% | 41 | 0.05 | 875 | -58 | -101 | 500 |
| Mauritius | 4,757 | 9% | 10% | 63 | 0.01 | 5,763 | -13 | -3 | 4,590 |
| Mexico | 6,333 | 9% | 10% | 3,344 | 0.51 | 6,862 | 0 | 0 | 6,333 |
| Moldova | 4,854 | 12% | 15% | 260 | 0.05 | 6,197 | -17 | -4 | 4,781 |
| Mongolia | 3,287 | 9% | 10% | 94 | 0.03 | 3,964 | 0 | 0 | 3,287 |
| Morocco | 2,609 | 12% | 15% | 1,605 | 0.52 | 3,600 | -37 | -14 | 2,591 |
| Mozambique | 276 | 11% | 15% | 274 | 0.82 | 400 | -70 | -271 | 251 |
| Namibia | 2,628 | 12% | 15% | 126 | 0.04 | 3,342 | -64 | -26 | 2,337 |
| Nepal | 480 | 8% | 10% | 226 | 0.41 | 640 | -15 | -33 | 471 |
| Nicaragua | 2,697 | 19% | 19% | 0 | 0.00 | 2,697 | -23 | -9 | 2,627 |
| Niger | 81 | 10% | 10% | 3 | 0.04 | 82 | 401 | 2,749 | 245 |
| Nigeria | 137 | 6% | 10% | 2,423 | 12.23 | 279 | 1,879 | 10,154 | 245 |
| Pakistan | 187 | 5% | 10% | 2,788 | 9.77 | 421 | 738 | 3,444 | 245 |
| Paraguay | 5,360 | 22% | 22% | 0 | 0.00 | 5,360 | -1 | 0 | 5,357 |
| Peru | 5,431 | 16% | 16% | 0 | 0.00 | 5,431 | 0 | 0 | 5,431 |
| Philippines | 1,658 | 9% | 10% | 1,272 | 0.71 | 1,930 | -20 | -12 | 1,654 |
| Rwanda | 520 | 13% | 15% | 60 | 0.11 | 594 | -164 | -393 | 357 |
| Senegal | 488 | 6% | 10% | 473 | 0.67 | 999 | -200 | -522 | 323 |
| Sierra Leone | 59 | 4% | 10% | 60 | 0.61 | 157 | 108 | 852 | 245 |
| South Africa | 3,206 | 13% | 15% | 3,041 | 0.85 | 3,966 | 0 | 0 | 3,206 |
| Sri Lanka | 1,149 | 7% | 10% | 425 | 0.31 | 1,647 | -170 | -162 | 985 |
| Sudan | 86 | 8% | 10% | 37 | 0.39 | 106 | 266 | 1,777 | 245 |
| Swaziland | 1,732 | 13% | 15% | 28 | 0.01 | 2,059 | -34 | -22 | 1,416 |
| Tajikistan | 595 | 8% | 10% | 94 | 0.13 | 840 | -53 | -100 | 485 |
| Tanzania | 276 | 8% | 10% | 294 | 0.93 | 361 | -107 | -412 | 250 |
| Thailand | 3,978 | 16% | 16% | 0 | 0.00 | 3,978 | -15 | -4 | 3,975 |
| The Gambia | 162 | 5% | 10% | 24 | 0.10 | 338 | 12 | 59 | 245 |
| Togo | 178 | 5% | 10% | 102 | 0.39 | 375 | 36 | 173 | 245 |
| Tunisia | 3,299 | 13% | 15% | 393 | 0.11 | 4,037 | -102 | -32 | 3,144 |
| Turkey | 7,957 | 10% | 10% | 804 | 0.10 | 8,174 | 0 | 0 | 7,957 |
| Turkmenistan | 1,822 | 9% | 10% | 84 | 0.04 | 2,153 | -6 | -3 | 1,802 |
| Uganda | 207 | 8% | 10% | 216 | 0.89 | 285 | 108 | 478 | 245 |
| Uzbekistan | 1,326 | 8% | 10% | 637 | 0.42 | 1,761 | -5 | -4 | 1,323 |
| Venezuela | 903 | 4% | 10% | 2,672 | 1.54 | 3,091 | 0 | 0 | 903 |
| Vietnam | 1,723 | 11% | 15% | 3,818 | 1.79 | 2,628 | -117 | -68 | 1,701 |
| Yemen | 88 | 9% | 10% | 28 | 0.30 | 105 | 238 | 1,580 | 245 |
| Zambia | 710 | 14% | 15% | 46 | 0.06 | 746 | -422 | -790 | 438 |
| Zimbabwe | 335 | 6% | 10% | 275 | 0.60 | 623 | -86 | -296 | 265 |

Note: Ukraine is excluded from the table as the WHO health expenditure database (GHED) has no data on the split of donor funding for 2022 or 2023.

**Table 2.** Estimated allocation, DALYs averted, and resulting marginal cost per DALY averted under an optimal allocation of the full development assistance for health (DAH) envelope, by country.

| Allocation of \$25.7 billion off-budget donor funding | | | |
| --- | --- | --- | --- |
| | Funding allocated (millions 2023 US\$) | DALYs averted by additional DAH (millions) | Marginal cost per DALY averted (2023 US\$) |
| Albania | 0 | 0 | 5,513 |
| Algeria | 0 | 0 | 2,512 |
| Argentina | 0 | 0 | 16,620 |
| Armenia | 0 | 0 | 2,234 |
| Azerbaijan | 0 | 0 | 1,751 |
| Bangladesh | 2,168 | 6 | 578 |
| Belarus | 0 | 0 | 5,673 |
| Belize | 0 | 0 | 3,939 |
| Benin | 289 | 1 | 578 |
| Bolivia | 0 | 0 | 3,001 |
| Botswana | 0 | 0 | 5,695 |
| Brazil | 0 | 0 | 7,195 |
| Burkina Faso | 524 | 1 | 578 |
| Burundi | 284 | 1 | 578 |
| Cambodia | 13 | 0 | 591 |
| Cameroon | 695 | 2 | 578 |
| Cape Verde | 0 | 0 | 3,414 |
| Chad | 514 | 1 | 578 |
| China | 0 | 0 | 8,766 |
| Colombia | 0 | 0 | 8,456 |
| Comoros | 15 | 0 | 586 |
| Congo | 89 | 0 | 578 |
| Costa Rica | 0 | 0 | 16,212 |
| Cote d'Ivoire | 376 | 1 | 578 |
| Democratic Republic of the Congo | 1,774 | 5 | 578 |
| Dominican Republic | 0 | 0 | 5,673 |
| Ecuador | 0 | 0 | 5,718 |
| Egypt | 0 | 0 | 985 |
| El Salvador | 0 | 0 | 5,230 |
| Eritrea | 139 | 0 | 578 |
| Ethiopia | 2,012 | 5 | 578 |
| Gabon | 0 | 0 | 2,343 |
| Georgia | 0 | 0 | 2,816 |
| Ghana | 0 | 0 | 647 |
| Guatemala | 0 | 0 | 2,387 |
| Guinea | 317 | 1 | 578 |
| Guinea-Bissau | 47 | 0 | 578 |
| Haiti | 255 | 1 | 578 |
| Honduras | 0 | 0 | 1,727 |
| India | 0 | 0 | 597 |
| Indonesia | 0 | 0 | 1,215 |
| Jamaica | 0 | 0 | 6,483 |
| Jordan | 0 | 0 | 3,874 |
| Kazakhstan | 0 | 0 | 6,324 |
| Kenya | 0 | 0 | 621 |
| Kyrgyzstan | 0 | 0 | 1,188 |
| Lebanon | 0 | 0 | 1,972 |
| Lesotho | 0 | 0 | 627 |
| Macedonia | 0 | 0 | 6,334 |
| Madagascar | 506 | 1 | 578 |
| Malawi | 388 | 1 | 578 |
| Malaysia | 0 | 0 | 5,133 |
| Mali | 569 | 1 | 578 |
| Mauritania | 29 | 0 | 598 |
| Mauritius | 0 | 0 | 4,590 |
| Mexico | 0 | 0 | 6,333 |
| Moldova | 0 | 0 | 4,781 |
| Mongolia | 0 | 0 | 3,287 |
| Morocco | 0 | 0 | 2,591 |
| Mozambique | 713 | 2 | 578 |
| Namibia | 0 | 0 | 2,337 |
| Nepal | 156 | 0 | 578 |
| Nicaragua | 0 | 0 | 2,627 |
| Niger | 687 | 2 | 578 |
| Nigeria | 5,061 | 13 | 578 |
| Pakistan | 3,753 | 10 | 578 |
| Paraguay | 0 | 0 | 5,357 |
| Peru | 0 | 0 | 5,431 |
| Philippines | 0 | 0 | 1,654 |
| Rwanda | 212 | 0 | 578 |
| Senegal | 288 | 1 | 578 |
| Sierra Leone | 160 | 0 | 578 |
| South Africa | 0 | 0 | 3,206 |
| Sri Lanka | 0 | 0 | 985 |
| Sudan | 471 | 1 | 578 |
| Swaziland | 0 | 0 | 1,416 |
| Tajikistan | 46 | 0 | 581 |
| Tanzania | 1,095 | 3 | 578 |
| Thailand | 0 | 0 | 3,975 |
| The Gambia | 42 | 0 | 578 |
| Togo | 161 | 0 | 578 |
| Tunisia | 0 | 0 | 3,144 |
| Turkey | 0 | 0 | 7,957 |
| Turkmenistan | 0 | 0 | 1,802 |
| Uganda | 842 | 2 | 578 |
| Uzbekistan | 0 | 0 | 1,323 |
| Venezuela | 0 | 0 | 903 |
| Vietnam | 0 | 0 | 1,701 |
| Yemen | 428 | 1 | 578 |
| Zambia | 251 | 1 | 599 |
| Zimbabwe | 321 | 1 | 578 |

Note: Ukraine is excluded from the table as the WHO health expenditure database (GHED) has no data on the split of donor funding for 2022 or 2023.

Among the 91 countries in scenario analyses GHE as a percent of GGE ranges from 2 to 26%, and 76 countries (84%) spend less than 15% of GGE on health. Increasing GHE to the nearest target of 10% or 15% would avert 145 million DALYs and require an increase in spending across countries of $392 billion. Results for increasing GHE as a percent of GGE to 15% as per the Abuja Declaration target in the 44 African Union countries in the sample are reported in the appendix.

According to the WHO GHED data 80 of 91 countries in our analysis are recipients of on-budget support from donor aid for health, totalling $11.2 billion. This aid, as currently allocated averts 45.3 million DALYs.^6^ Reallocating this pot of money according to where it would have the greatest health benefit would avert 75.3 million DALYs, a gain of 30.0 million DALYs from targeting alone, with no change in total resources. This reallocation involves substantial redistribution with 57 countries losing their on-budget funding entirely, and four countries receiving reduced funding, as it is redirected to 22 countries where it generates more health. Concentrating this limited pot where it generates the greatest health benefit substantially increases its impact, though at the cost of withdrawing support from a majority of currently-funded countries.

Separately, we consider the potential of the full DAH envelope, which, unlike on-budget support, includes off-budget, NGO, and philanthropic funding not currently tracked at the country level. A further 66.1 million DALYs averted are attributable to the additional approximately $25.7 billion in off-budget, NGO, and philanthropic resources, reflecting the additional scale of resources available once the full DAH envelope, rather than on-budget support alone, is considered.

## Discussion

This paper provides updated marginal cost per DALY averted estimates for 92 LMIC. For most countries, the estimated marginal cost per DALY averted is higher than in the previous analysis of cross-country data from 2018. However, as with the previous analysis, this paper finds that most estimates of cost per DALY averted are below 1x GDP per capita, reinforcing the earlier finding that applying GDP per capita based cost-effectiveness thresholds to funding and prioritisation decisions risks reducing overall population health.

Patterns of health expenditure, disease burden, and demographic structures in countries have changed between the studies. Each of these influences the cost per DALY averted. All else equal, a decrease in mortality would result in a higher cost per DALY averted; because baseline mortality is lower, the same percentage change in mortality for a given percentage change in expenditure is lower. However, these factors interact, and the implied cost per DALY averted ultimately reflects the opportunity cost of expenditure within the mix of funded services (31).

The increase in cost per DALY averted in this analysis across most countries is largely driven by increases in GHE over time, reflecting movement along a concave health production function in which, all else equal, marginal returns diminish as spending increases (10,32). In a minority of countries, declines in GHE between the periods of analysis or revisions to historical expenditure data contributed to lower estimated costs per DALY averted. Brazil and Lebanon are exceptions: their estimates fell even though their revised 2014 GHE was not lower than the value used in the 2018 analysis. In Lebanon, GHE per capita fell sharply between 2014 and 2023, from $271 to $103 using current data, during the country’s economic crisis (33). In Brazil, GHE per capita changed little over the period (a fall of 2% using current data), while DALYs increased by 17%, mainly due to an increase in YLDs. Where this follows revisions to historical expenditure data, this suggests that GHE is in fact more productive than previously estimated.

These updated estimates provide a basis for countries to assess not only whether new interventions generate more health than they displace, but the magnitude of the gains or losses (5,6). In doing so, they strengthen locally led, evidence-informed priority-setting processes. Beyond informing domestic priority setting, these estimates have direct implications for how scarce donor resources should be allocated across countries.

If the objective of donors is to maximise global health gains, then resources should be directed to settings where the marginal productivity of health expenditure is highest, that is, where the cost per DALY averted is lowest. The current best estimate of on-budget support as currently allocated averts 45.3 million DALYs, while allocating it in line with country-specific opportunity costs would avert approximately 75.3 million DALYs, a substantial gain achieved purely by moving funding from countries where it generates less health benefit to countries where it generates more, without any increase in total resources. This reflects that the health returns to spending vary substantially across countries, and that a donor strategy which explicitly accounts for these differences can achieve markedly greater impact at the margin. However, this gain is not without cost: achieving it requires withdrawing on-budget support entirely from 57 of the 80 currently-funded countries, raising practical questions about transition costs and political feasibility that we discuss further below.

Beyond this targeting effect, the scale of resources considered also matters substantially. In addition to the 75.3 million DALYs attributable to optimal on-budget targeting (compared to 45.3 million from on-budget support as currently allocated), a further 66.1 million is attributable to the additional resources available once the fuller DAH envelope is considered. Together these results indicate that both better targeting of existing on-budget aid and mobilising the broader resource envelope toward country-specific needs offer meaningful, and largely additive, opportunities for impact.

Raising domestic spending to 15%, or even 10%, of general government expenditure would require significant increases in domestic spending on health. That the spend required to reach this target far outstrips current DAH underscores the challenge of doing so. In the context of constrained and declining donor funding (34), it is increasingly important for countries to strengthen domestic resource mobilisation for health, while ensuring that both domestic and external funds are allocated where they generate the greatest health gains.

The tiered scenario results are heavily influenced by a single country. China accounts for $280 billion of the $392 billion in additional expenditure but only 25 million of the 145 million DALYs averted. Excluding China, raising GHE to the tiered targets would require $112 billion and avert 120 million DALYs. Most of the health gain comes from the first step: raising the 49 countries spending less than 10% of general government expenditure on health to 10% would cost $94 billion and avert 112 million DALYs (around $840 per DALY averted), whereas raising the 27 countries spending between 10% and 15% to 15% would cost $298 billion and avert 33 million DALYs (around $8,900 per DALY averted). Increasing the share of government budgets allocated to health on this scale would also displace spending in other sectors, such as education, transport or environmental protection, some of which also improves health. These cross-sectoral opportunity costs may not be captured depending on what is captured in the estimated elasticity.

In the scenario analyses additional financing, whether financed domestically or through donors, is treated as augmenting domestic health spending without altering the underlying health production function. This implies that domestic and donor-financed expenditure are considered interchangeable with GHE with respect to their productivity at the margin. In practice, donor funding may interact with domestic spending through mechanisms including crowd-out, co-financing requirements, or institutional strengthening that affects the productivity of health systems. The estimates presented here should therefore be interpreted as approximating the health gains from additional resources entering the health system under the assumption that the productivity of expenditure is invariant to its financing source. Other papers have assessed different types of investments in healthcare and further research in this area would be valuable (35).

The donor allocation results should be interpreted as a theoretical optimum rather than a directly implementable policy. They assume that recipient health systems can absorb additional resources at the estimated marginal productivity, and that funding can be withdrawn from some countries and redirected to others without transition costs. In practice, governance, institutional quality, health system capacity and absorptive capacity differ across countries and may influence the health gains realised, particularly where allocations are large relative to current spending. Under the full-envelope allocation, funding exceeds current GHE in 25 of the 37 recipient countries and is more than double current GHE in 15, reaching around five and a half times current GHE in Sierra Leone. Similarly, withdrawing on-budget support from 57 of the 80 currently funded countries would involve transition costs and is unlikely to be politically feasible over a short period. The value of the exercise is therefore not to suggest that all DAH could immediately be redistributed in this way, but to quantify an estimate of the health that could be achieved, and hence the opportunity cost, in health forgone, of departures from that allocation.

Recent analysis has argued that achieving value for money in global development requires moving beyond short-term project metrics toward strengthening national priority-setting systems, supporting domestic co-financing, consolidating fragmented funding streams, and aligning donor investments with recipient government priorities (36). A central recommendation is that aid should increasingly focus on helping countries use their own resources more effectively, thereby generating durable improvements in health system performance that outlast direct donor involvement. Under a New Compact model of global health financing, where countries assume greater responsibility for financing core services and donors provide coordinated support at the margin, credible opportunity cost estimates become a foundational input (37). Investments in common goods that strengthen health systems may yield significant health gains, and quantifying their potential net health effects across countries requires estimates of the costs, expected benefits and the opportunity cost of such investments in each country (7).

Allocating donor funding according to the marginal cost per DALY averted could, in principle, create perverse incentives. Because the estimated cost per DALY averted is lower where GHE is lower, a country that reduced its domestic health spending would appear more productive at the margin and so attract more donor funding. Similarly, all else equal, countries with a higher burden of disease would receive more funding. The purpose of this analysis is to illustrate where the returns to additional spending are greatest given current estimates, not to propose a mechanical allocation rule. Incentives of this kind could be anticipated in the design of any allocation mechanism, for example by making donor funding conditional on countries not reducing GHE without strong justification, consistent with the domestic-first principle of the New Compact. While it is unlikely that a country would seek to increase its burden of disease to attract funding, related issues could be anticipated in the same way.

Such an approach has been proposed in the UK where recent reductions in bilateral aid budgets, including those of the UK Foreign, Commonwealth & Development Office (FCDO), have intensified scrutiny of how limited development resources are allocated (36,38). UK parliamentary and policy discussions on value for money have emphasised that, in a constrained fiscal environment, aid must demonstrably maximise impact while supporting long-term sustainability (39). In this context, calls for a renewed focus on efficiency, domestic resource mobilisation, and institutional strengthening are particularly salient.

These issues are particularly topical given changes in the global health architecture. Under the bilateral health agreements signed as part of the United States’ America First Global Health Strategy, partner countries commit to increasing domestic co-investment in health as US assistance declines; across the 36 countries with signed agreements, co-investment is projected to make up 39% of the $26.7 billion in total announced funding over 2026 to 2030 (40). Other global health initiatives, such as the Global Fund, also set domestic co-financing requirements (41). It is important to distinguish between increasing domestic health expenditure and requiring additional domestic expenditure to be earmarked for donor-selected programmes. Where co-financing is tied to specific vertical programmes, it determines not only how much domestic money is spent on health but what it is spent on. If the earmarked programmes have a higher cost per DALY averted than the services that would otherwise be funded at the margin, the result is a net loss of health, borne by the domestic budget as well as the donor. The estimates presented here provide a basis for quantifying such losses and for assessing whether co-financing requirements are consistent with the health that would be forgone, which is a topical area for further work.

In addition to informing allocation across countries, country-specific estimates of the marginal cost per DALY averted can inform value-based pricing within pooled procurement arrangements. Under pooled mechanisms, for example for vaccines, antiretrovirals, or other high-impact interventions, suppliers negotiate prices across multiple participating countries (42). Aligning such negotiations with country-specific opportunity cost estimates enables pooled procurement bodies to define price ceilings consistent with the health that would otherwise be displaced within each health system (43). In this way, pooled procurement can move beyond securing lower prices in absolute terms toward ensuring that negotiated prices are consistent with net health gains across countries.

By providing updated, country-specific estimates of the marginal productivity of health expenditure, this study equips both donors and governments with empirical evidence to inform assessments of the health value of additional spending, whether domestic or external. In a context of shrinking aid envelopes, such estimates become even more important as they help to ensure that scarce donor resources are directed to settings where marginal health gains are greatest, while also strengthening domestic priority-setting institutions to improve the efficiency of nationally financed spending.

Several limitations warrant consideration. First, we retain previously estimated elasticity parameters from Ochalek et al (2018) and update the underlying country-level inputs. If the relationship between expenditure and health outcomes has structurally changed over time, our estimates may not fully capture this. This should be a priority for future research. Second, scenario modelling is based on one of the four DALY measures for tractability, though results are broadly representative across specifications. Third, the analysis excludes out-of-pocket expenditure and focuses on government and compulsory contributory schemes, consistent with the underlying framework. The relationship between GHE and out-of-pocket expenditure is discussed elsewhere (forthcoming). Fourth, estimates are derived at the national level and do not capture within-country heterogeneity or political feasibility constraints.

The first of these limitations warrants particular attention. Retaining a single set of elasticities assumes that the proportional responsiveness of health outcomes to GHE is the same across countries and has not changed since they were estimated. Updating expenditure, disease burden and population captures movement along the health production function, including diminishing returns as spending rises, but not shifts in the function itself. Such shifts are plausible given changes in epidemiology, service delivery and the composition of spending, and shocks like the Covid-19 pandemic (31). The relationship between spending and health is also likely to differ between low-and upper-middle-income countries and to be mediated by factors such as how health systems are financed and the strength of governance and institutions. The direction of any resulting bias is unclear. A clearer research agenda is needed to address this, including re-estimating the elasticities using more recent data, and allowing elasticities to vary by health system characteristics such as financing arrangements and governance. Furthermore, differences between the estimates presented here and those from 2018 may reflect not only changes in disease burden and population, but also changes in GBD methods, disease models and input data between releases, as well as the revisions to expenditure data discussed above.

Uncertainty is only partially reflected in the results. The estimates are point estimates, and uncertainty in the elasticities, the GBD estimates of disease burden and the expenditure data is not propagated through the analysis; the range across the four DALY measures (Figure 1) indicates sensitivity to the choice of DALY measure. A probabilistic sensitivity analysis would provide a fuller characterisation of uncertainty and should be a priority for future updates.

These limitations, however, do not alter the central implication: opportunity costs differ substantially across countries and evolve over time, with important consequences for both domestic and donor allocation decisions. Regularly updating opportunity cost estimates is essential, particularly so in a context where both domestic budgets and donor financing are volatile. Static thresholds based on historical data risk misrepresenting current trade-offs and undermining efficient priority setting.

## Conclusion

In a context of tightening fiscal space and declining external assistance, the opportunity cost of misallocation of resources for healthcare is more relevant than ever. Updated estimates of the marginal cost per DALY averted across countries can inform decisions around how to allocate government resources for health, as well as informing domestic-first financing and consolidated supplementary aid under a New Compact for global health financing. Aligning donor support with country-specific opportunity costs can maximise health gains.

## Data Availability

All data analysed in this study are publicly available, and the country-level health opportunity cost estimates generated in this study are provided in the Additional files. The analysis code and derived datasets are available from the author on reasonable request.

https://apps.who.int/nha/database

https://ghdx.healthdata.org/

https://datahelpdesk.worldbank.org/knowledgebase/articles/906519-world-bank-country-and-lending-groups

## List of Abbreviations

DAC: Development Assistance Committee (of the OECD)
DAH: development assistance for health
DALY: disability-adjusted life year
EQ-5D: EuroQol 5-Dimension
FCDO: Foreign, Commonwealth & Development Office
GBD: Global Burden of Disease
GDP: gross domestic product
GGE: general government expenditure
GHDx: Global Health Data Exchange
GHE: government health expenditure
GHED: Global Health Expenditure Database
IHME: Institute for Health Metrics and Evaluation
IRS: Internal Revenue Service
LMIC: low- and middle-income countries
NGO: non-governmental organisation
ODA: official development assistance
OECD: Organisation for Economic Co-operation and Development
OOP: out-of-pocket
QALY: quality-adjusted life year
UK: United Kingdom
US: United States
WHO: World Health Organization
YLD: years lived with disability
YLL: years of life lost

## Declarations

### Ethics approval and consent to participate

Not applicable. This study uses publicly available, aggregate country-level data and did not involve human participants.

### Consent for publication

Not applicable.

### Availability of data and materials

All data used in this study are publicly available from the WHO Global Health Expenditure Database, the Global Burden of Disease Study 2023 (via the Global Health Data Exchange), the World Bank World Development Indicators, and the IHME Development Assistance for Health Database.

### Competing interests

JO carried out this work as a paid consultant to the Center for Global Development. No other competing interests.

### Funding

This study was supported by the Center for Global Development, through the Foreign Commonwealth and Development Office, project code: 301528-401, and Coefficient Giving.

### Authors’ contributions

JO designed the study with input from the Center for Global Development, carried out the data analysis and interpretation, and drafted and revised the manuscript. The author read and approved the final manuscript.

This work was carried out as private consultancy by the author. Although the author is otherwise employed by the University of York, the work and recommendations produced in this paper do not imply an endorsement or commitment from that university.

## Acknowledgements

The author gratefully acknowledges comments from Pete Baker, Rachel Bonnifield, Anastassia Demeshko and Adrian Gheorghe on an earlier draft, comments from anonymous reviewers on an earlier submitted version, and discussions with James Lomas.

## Footnotes

1 This includes DAH for countries excluded from our sample.

2 Health expenditure managed through government schemes and compulsory contributory schemes (HF.1) includes: transfers from government domestic revenue (allocated to health purposes); transfers distributed by government from foreign origin; social insurance contributions; compulsory prepayment (other, and unspecified, than FS.3); and unspecified revenues of health care financing schemes (n.e.c.).

3 Venezuela was not classified in the most recent World Bank Group income classifications for 2023, so we use the most recent classification.

4 Five countries that were included in the 2018 analysis are excluded from this analysis as they have moved out of the LMIC category into the high-income country category: Bulgaria, Guyana, Panama, Romania, Russia

5 Eritrea is excluded from this calculation and the figure, as there was no data for GDP per capita for any year from 2015 to current.

6 Recent data on the split between on- and off-budget support is not available for Ukraine.

## Notes

### Competing Interest Statement

The authors have declared no competing interest.

### Summary of Updates

This version has been revised following peer review. The main changes are: 1.The domestic financing scenario now uses a tiered target. Countries spending below 10 percent of general government expenditure on health move to 10 percent, and those between 10 and 15 percent move to 15 percent. The Abuja Declaration is used only to justify the upper bound, and the separate African Union analysis has been removed. 2.The DAH analysis is now split into two parts: reallocation of existing on-budget support (11.2 billion US dollars) to maximise health, and optimal allocation of the full DAH envelope. 3.Development assistance for health (DAH) totals from the IHME Financing Global Health database now exclude double-counted flows and unallocated, global and administrative categories, giving a total of 36.9 billion US dollars in 2023. 4.Data source attribution has been corrected to the WHO Global Health Expenditure Database. 5.Correction of a data error in the population calculation, where death counts for ages 5 to 9 and 10 to 14 had been copied from the under 5 age group. This overstated population, and therefore total health expenditure, in some countries, particularly in sub-Saharan Africa. All estimates, tables and figures have been updated. 6.The Discussion has been expanded to address reviewer comments, including the relationship between the domestic scenario and the New Compact, the treatment of philanthropic funding, and limitations of the approach. 7.All monetary values are now consistently reported in constant 2023 US dollars.

