## Supplemental Tables for "Updated Health Opportunity Cost Estimates for 92 Low- and Middle-Income Countries: Implications for Global Health Financing and Donor Allocation"

Appendix Table 1 below presents the cost per DALY averted estimate from each DALY measure calculated and for average across the four measures.

**Appendix Table 1. Cost per DALY averted (2023 US$) for 92 LMIC**

| Country | **Cost per DALY 1** | **Cost per DALY 2** | **Cost per DALY 3** | **Cost per DALY 4** | **Cost per average DALYs averted** |
|---|---|---|---|---|---|
| Albania | $6,979 | $4,286 | $7,348 | $5,513 | $5,763 |
| Algeria | $2,561 | $2,095 | $3,158 | $2,512 | $2,528 |
| Argentina | $21,684 | $14,845 | $21,878 | $16,852 | $18,304 |
| Armenia | $2,492 | $1,887 | $2,938 | $2,256 | $2,333 |
| Azerbaijan | $1,668 | $1,457 | $2,032 | $1,770 | $1,707 |
| Bangladesh | $133 | $113 | $164 | $143 | $136 |
| Belarus | $6,687 | $4,556 | $6,686 | $5,673 | $5,756 |
| Belize | $6,118 | $3,521 | $4,864 | $3,998 | $4,429 |
| Benin | $225 | $190 | $212 | $232 | $214 |
| Bolivia | $3,264 | $2,536 | $3,408 | $3,011 | $3,016 |
| Botswana | $6,956 | $4,613 | $6,352 | $5,914 | $5,822 |
| Brazil | $8,712 | $6,274 | $9,111 | $7,196 | $7,650 |
| Burkina Faso | $375 | $349 | $384 | $410 | $378 |
| Burundi | $88 | $78 | $82 | $93 | $85 |
| Cambodia | $834 | $528 | $715 | $651 | $664 |
| Cameroon | $200 | $183 | $200 | $218 | $200 |
| Cape Verde | $4,151 | $3,050 | $4,686 | $3,908 | $3,853 |
| Chad | $122 | $120 | $126 | $151 | $128 |
| China | $11,061 | $7,509 | $11,697 | $8,766 | $9,453 |
| Colombia | $10,497 | $7,464 | $11,072 | $8,457 | $9,137 |
| Comoros | $508 | $505 | $651 | $623 | $564 |
| Congo | $450 | $376 | $450 | $456 | $430 |
| Costa Rica | $23,266 | $14,938 | $22,688 | $16,215 | $18,547 |
| Cote d'Ivoire | $446 | $403 | $439 | $478 | $440 |
| Democratic Republic of the Congo | $80 | $72 | $85 | $93 | $82 |
| Dominican Republic | $7,842 | $4,918 | $6,728 | $5,707 | $6,110 |
| Ecuador | $6,144 | $4,821 | $6,657 | $5,721 | $5,754 |
| Egypt | $1,102 | $904 | $1,332 | $1,078 | $1,084 |
| El Salvador | $6,227 | $4,333 | $6,149 | $5,240 | $5,370 |
| Eritrea | $137 | $115 | $127 | $139 | $129 |
| Ethiopia | $150 | $132 | $154 | $166 | $149 |
| Gabon | $2,565 | $1,928 | $2,539 | $2,472 | $2,344 |
| Georgia | $3,165 | $2,258 | $3,398 | $2,842 | $2,847 |
| Ghana | $663 | $549 | $648 | $658 | $626 |
| Guatemala | $2,667 | $2,119 | $2,953 | $2,501 | $2,523 |
| Guinea | $171 | $158 | $179 | $201 | $176 |
| Guinea-Bissau | $223 | $200 | $227 | $247 | $223 |
| Haiti | $160 | $105 | $119 | $123 | $123 |

| | | | | | |
|---|---|---|---|---|---|
| Honduras | $2,975 | $2,077 | $2,893 | $2,190 | $2,469 |
| India | $591 | $509 | $741 | $629 | $607 |
| Indonesia | $1,307 | $983 | $1,307 | $1,223 | $1,188 |
| Jamaica | $6,396 | $5,206 | $7,786 | $6,533 | $6,350 |
| Jordan | $6,013 | $4,452 | $6,117 | $4,161 | $5,033 |
| Kazakhstan | $7,639 | $5,262 | $7,605 | $6,339 | $6,555 |
| Kenya | $684 | $559 | $745 | $729 | $671 |
| Kyrgyzstan | $1,489 | $1,018 | $1,458 | $1,190 | $1,258 |
| Lebanon | $1,857 | $1,604 | $2,741 | $2,050 | $1,985 |
| Lesotho | $847 | $661 | $823 | $875 | $792 |
| Macedonia | $10,132 | $5,190 | $8,062 | $6,334 | $6,978 |
| Madagascar | $87 | $78 | $90 | $98 | $88 |
| Malawi | $211 | $181 | $208 | $224 | $205 |
| Malaysia | $6,371 | $4,153 | $6,422 | $5,133 | $5,346 |
| Mali | $120 | $115 | $120 | $137 | $123 |
| Mauritania | $602 | $542 | $699 | $698 | $628 |
| Mauritius | $5,109 | $3,945 | $5,524 | $4,757 | $4,760 |
| Mexico | $8,043 | $5,947 | $8,077 | $6,333 | $6,966 |
| Moldova | $5,498 | $3,858 | $5,667 | $4,854 | $4,857 |
| Mongolia | $3,868 | $2,727 | $3,772 | $3,287 | $3,348 |
| Morocco | $2,805 | $2,098 | $3,179 | $2,609 | $2,613 |
| Mozambique | $275 | $225 | $259 | $276 | $257 |
| Namibia | $3,665 | $2,158 | $2,564 | $2,628 | $2,655 |
| Nepal | $491 | $383 | $539 | $480 | $466 |
| Nicaragua | $5,321 | $2,492 | $3,505 | $2,697 | $3,212 |
| Niger | $65 | $65 | $67 | $81 | $69 |
| Nigeria | $114 | $112 | $123 | $137 | $121 |
| Pakistan | $164 | $154 | $191 | $187 | $172 |
| Paraguay | $8,103 | $4,439 | $6,538 | $5,360 | $5,812 |
| Peru | $6,227 | $4,769 | $6,682 | $5,431 | $5,681 |
| Philippines | $1,592 | $1,291 | $1,831 | $1,658 | $1,568 |
| Rwanda | $540 | $425 | $495 | $520 | $491 |
| Senegal | $532 | $411 | $491 | $488 | $476 |
| Sierra Leone | $55 | $49 | $52 | $59 | $54 |
| South Africa | $3,631 | $2,700 | $3,332 | $3,206 | $3,180 |
| Sri Lanka | $1,185 | $932 | $1,377 | $1,149 | $1,138 |
| Sudan | $82 | $65 | $94 | $86 | $80 |
| Swaziland | $2,060 | $1,299 | $1,623 | $1,732 | $1,634 |
| Tajikistan | $570 | $502 | $687 | $595 | $581 |
| Tanzania | $253 | $226 | $274 | $276 | $256 |
| Thailand | $4,947 | $3,198 | $4,478 | $3,978 | $4,043 |
| The Gambia | $156 | $137 | $157 | $162 | $152 |
| Togo | $178 | $149 | $171 | $178 | $168 |
| Tunisia | $3,309 | $2,784 | $4,228 | $3,299 | $3,331 |
| Turkey | $8,774 | $6,866 | $10,392 | $7,957 | $8,307 |
| Turkmenistan | $1,771 | $1,443 | $1,957 | $1,822 | $1,726 |
| Uganda | $197 | $166 | $191 | $207 | $189 |
| Uzbekistan | $1,180 | $1,082 | $1,457 | $1,326 | $1,245 |
| Venezuela | $913 | $696 | $1,044 | $903 | $870 |
| Vietnam | $2,020 | $1,386 | $1,946 | $1,723 | $1,731 |
| Yemen | $89 | $83 | $106 | $88 | $91 |
| Zambia | $883 | $600 | $682 | $710 | $705 |

| | | | | | |
|---|---|---|---|---|---|
| Zimbabwe | $348 | $263 | $318 | $335 | $313 |

Appendix Table 2 below reports the potential health effects of increasing GHE as a percent of GGE to 15% in all African Union member states included in our analysis. There is a potential to avert 61 million DALYs, but at a cost of 35.8 billion in additional expenditure on health. Since this comes from GGE, it would require either reallocating spending from other sectors or increasing total government expenditure through taxation or borrowing. Reallocation would displace spending in areas such as education, social protection and infrastructure, which themselves contribute to health and wellbeing, so the net health gains are likely to be smaller than those reported here. The scale of the additional expenditure is also notable: it is almost equal to the entire 2023 DAH envelope ($36.9 billion, 2023 US$). For these reasons, the main analysis uses a tiered target, with the Abuja target of 15% serving as its upper bound.

**Appendix Table 2. Increasing GHE to 15% of GGE in AU countries**

| | Increase of GHE as a % of GHE to 15% | | | |
|---|---|---|---|---|
| **Country** | **Current GHE as a % of GGE** | **Additional expenditure required (millions 2023 US$)** | **DALYs averted by additional expenditure (millions)** | **Marginal cost per DALY averted (2023 US$)** |
| Algeria | 6% | 7,909 | 1.78 | 4,513 |
| Benin | 7% | 304 | 0.81 | 355 |
| Botswana | 15% | 0 | 0.00 | 5,914 |
| Burkina Faso | 13% | 131 | 0.29 | 492 |
| Burundi | 10% | 64 | 0.53 | 94 |
| Cameroon | 6% | 901 | 2.31 | 399 |
| Cape Verde | 16% | 0 | 0.00 | 3,908 |
| Chad | 10% | 145 | 0.77 | 233 |
| Comoros | 13% | 5 | 0.01 | 742 |
| Congo | 7% | 234 | 0.31 | 706 |
| Cote d'Ivoire | 7% | 1,410 | 1.88 | 693 |
| Democratic Republic of the Congo | 7% | 781 | 5.41 | 132 |
| Egypt | 8% | 5,133 | 3.18 | 1,430 |
| Eritrea | 4% | 206 | 0.66 | 379 |
| Ethiopia | 9% | 877 | 3.92 | 182 |
| Gabon | 10% | 162 | 0.05 | 3,985 |
| Ghana | 10% | 705 | 0.84 | 1,047 |
| Guinea | 7% | 279 | 0.88 | 296 |
| Guinea-Bissau | 10% | 21 | 0.07 | 386 |
| Kenya | 10% | 1,191 | 1.25 | 745 |
| Lesotho | 17% | 0 | 0.00 | 875 |
| Madagascar | 8% | 192 | 1.34 | 125 |
| Malawi | 10% | 152 | 0.54 | 347 |
| Mali | 6% | 466 | 1.96 | 237 |
| Mauritania | 8% | 164 | 0.16 | 875 |
| Mauritius | 9% | 275 | 0.04 | 5,763 |

| | | | | |
|---|---|---|---|---|
| Morocco | 12% | 1,605 | 0.52 | 3,600 |
| Mozambique | 11% | 274 | 0.82 | 400 |
| Namibia | 12% | 126 | 0.04 | 3,342 |
| Niger | 10% | 139 | 1.32 | 82 |
| Nigeria | 6% | 5,156 | 19.85 | 279 |
| Rwanda | 13% | 60 | 0.11 | 594 |
| Senegal | 6% | 1,004 | 1.08 | 999 |
| Sierra Leone | 4% | 114 | 0.87 | 157 |
| South Africa | 13% | 3,041 | 0.85 | 3,966 |
| Sudan | 8% | 152 | 1.23 | 106 |
| Swaziland | 13% | 28 | 0.01 | 2,059 |
| Tanzania | 8% | 1,027 | 2.50 | 361 |
| The Gambia | 5% | 51 | 0.16 | 338 |
| Togo | 5% | 214 | 0.62 | 375 |
| Tunisia | 13% | 393 | 0.11 | 4,037 |
| Uganda | 8% | 680 | 2.15 | 285 |
| Zambia | 14% | 46 | 0.06 | 746 |
| Zimbabwe | 6% | 619 | 1.02 | 623 |
